# Tuberculosis in households with infectious cases in Kampala city: Harnessing health data science for new insights on an ancient disease with persistent, unresolved problems (DS-IAFRICA-TB) Study Protocol

**DOI:** 10.64898/2026.04.23.26351571

**Authors:** Emmanuel Nasinghe, Denis Musinguzi, Mercy Takuwa, Rogers Kamulegeya, Rose Nabatanzi, Sharon Namiiro, Cosmas Mwikirize, Andrew Katumba, Willy Ssengooba, Florence N. Kivunike, Joyce Nakatumba-Nabende, David Patrick Kateete

## Abstract

Tuberculosis (TB) is prevalent in Uganda and overlaps with a high rate of HIV/TB coinfection. While nearly all hospital-based TB cases in Kampala, the capital of Uganda, show clear TB symptoms, 30% or more of undiagnosed TB cases found through active screening are asymptomatic. Additionally, the host risk factors for TB in Kampala cannot be distinguished from environmental risk factors. These TB-specific challenges are just part of the complexity, especially in areas with high HIV/AIDS burden. Data science techniques, especially Artificial Intelligence (AI) and Machine Learning (ML) algorithms, could help untangle this complexity by identifying factors related to the host, pathogen, and environment, which are difficult to explain or predict with traditional/conventional methods. In this project, we will use health data science approaches (AI/ML) to identify factors driving TB transmission within households and reasons for anti-TB treatment failure. We will utilize the computational resources at Makerere University and available demographic, clinical, and laboratory data from TB patients and their contacts to develop AI and ML algorithms. These will aim to: (1) identify patients at baseline (month 0) unlikely to convert their sputum or culture results by months 2 and 5, thus at risk of failing TB treatment; (2) identify household contacts of TB cases who are at risk of developing TB disease, as well as contacts who may resist TB infection despite repeated exposure to *M. tuberculosis*. Achieving these objectives will provide evidence that data science methods are effective for early detection of potential TB cases and high-risk patients, thereby helping to reduce TB transmission in the community. The study protocol received approval from the School of Biomedical Sciences IRB, protocol number SBS-2023-495.

## Introduction

Tuberculosis (TB) remains a major global public health concern, ranking as one of the deadliest infectious diseases alongside HIV/AIDS and malaria (1). It’s an ancient disease, as old as humankind, that still results in over a million deaths each year around the world (2). In 2024, about 8.3 million people were newly diagnosed and officially recorded with TB, slightly up from 8.2 million cases in 2023 (3).

In Uganda, a low-income country (LIC) with roughly 50 million residents, nearly 1 million new TB cases have been reported since 2010. The TB incidence has overtaken HIV in the country (4); in 2024, 197 new TB cases per 100,000 people were reported, including 65 per 100,000 among those HIV-positive (3). That year, Uganda was among the top 30 countries with the highest TB burden, overlapping with areas heavily impacted by TB/HIV coinfection (3).

*Mycobacterium tuberculosis*, the main causative agent of TB in humans, is among the most successful intracellular pathogens due to its high infectivity, widespread distribution (5), and capacity to adapt and evolve alongside humans (6). People are usually exposed to aerosols from infected individuals, which can trigger immune responses that lead to various clinical outcomes (5, 7). On one end is “TB resistance,” where the innate immune system clears bacteria from the respiratory mucosa, preventing infection and/or disease. On the other end is “active TB,” featuring symptoms similar to pneumonia—such as a persistent cough with bacteria-laden sputum—along with fever, systemic inflammation, and lung damage visible on X-ray (5, 8). Active TB can first appear as primary active TB after infection or emerge later as post-primary, reactivated TB. Between resistance and active disease, there is TB infection with no symptoms (sometimes referred to as latent TB), and asymptomatic active TB (referred to as ‘subclinical TB’) (5, 9, 10). These conditions exist on a spectrum, ranging from completely asymptomatic cases to those with subtle symptoms that might go unnoticed or be difficult to detect during screening (7, 11). Note that ‘subclinical TB’ can oscillate between ‘TB infection’, with no symptoms, and overt TB disease. Moreover, it has recently been confirmed that subclinical TB is transmissible despite the absence of symptoms (or failure to detect symptoms), highlighting the need for targeted control measures (12, 13).

Therefore, despite progress in TB control and immune protection, significant challenges persist. These include, in addition to the TB clinical conundrum described above, our failure to understand why some individuals can completely clear *M. tuberculosis* after infection, while others experience reactivation, and why only a small fraction of those with TB infection with no symptoms (i.e., around 15%) develop into active TB (5, 14). Although HIV-related CD4+ T cell decline increases reactivation risk, the precise mechanisms by which these cells influence control or failure are not fully known. Interestingly, TB reactivation during HIV isn’t solely linked to CD4+ cell loss, as some HIV-positive people develop TB before notable CD4+ decline. In fact, many HIV-infected individuals in Uganda and in other African countries can resist *M. tuberculosis* infection despite repeated exposures (15). Additionally, the full spectrum of TB infection and disease states is poorly understood in the context of TB/HIV co-infection.

Taken together, addressing TB in high-burden, high-HIV-prevalence LICs necessitates new strategies and multidisciplinary teams that combine clinical, biomedical, epidemiological, and computational expertise, leveraging big data and data-driven approaches in Africa. Therefore, this protocol aims to employ data-driven methods to tackle the complexity of TB phenotypes in Kampala, Uganda, an LIC setting heavily burdened by HIV/TB.

## Materials and Methods

### Aims

This protocol has two aims;

**Aim 1:** Develop Machine Learning (ML) models to predict, at baseline (month 0), which TB patients would not sputum/culture convert at months 2 and 5, and are therefore at risk of failing treatment with anti-tuberculous drugs.

**Aim 2:** Develop ML models to identify contacts of index-TB cases that are at risk of developing household TB disease, and predict contacts who could be resistant to TB infection despite persistent and/or multiple exposure to *M. tuberculosis* in a household.

### Study Design

This cross-sectional study will utilize existing de-identified TB datasets (secondary data). While the original studies included follow-up over time, our analysis adopts a cross-sectional approach, extracting and examining data at predefined time points for each participant.

### Study Setting

The study will be conducted in Kampala city. The datasets will be collected from research projects conducted at Kampala’s major TB diagnostic and treatment hospitals and centers, as well as from surrounding communities where index TB patients and their household contacts reside. Kampala has slum dwellings in each of its five divisions; such crowding and poor housing conditions in these areas contribute to a high burden of TB in Kampala District, which accounts for 25% of all TB cases in the country (15).

### Sample Size

Data science approaches rely on access to large, complex datasets to train and develop AI and ML algorithms. Therefore, we will absorb all available and/or usable datasets from the studies described above. We anticipate getting records from one thousand five hundred participants (1,500) across 3 protocols.

### Sampling criteria

The individuals and datasets to be included in the study will be selected through purposive sampling.

## Inclusion Criteria and Exclusion Criteria

### Inclusion criteria

We will include studies that meet the following conditions:

Study focus: Research studies that have collected data on the TB disease spectrum, including: TB incidence, disease state or severity, disease progression, and response to treatment

Study population: Studies that have recruited index TB patients and their household contacts, with clearly documented contact relationships.

Time period: Studies conducted or published from 1980 to the present. This period coincides with the advent and growing use of sequencing technologies in biological research.

Data availability: Studies that have stored their data in electronic formats or whose data is digitizable, enabling extraction and analysis.

### Exclusion criteria

Studies that do not include relevant TB data, such as details on the disease spectrum, progression, or treatment outcomes.

Studies that do not recruit index household contacts, including those focusing solely on community-level TB data, and those without defined contact structures.

Studies with non-electronic or inaccessible data, where raw data cannot be retrieved or verified.

## Study Variables

The Variables for Aim 1 are: demographic data, clinical history, laboratory findings, treatment-related variables, imaging results, symptoms and severity, genetic factors, compliance and adherence, healthcare system variables, microbiological data, response to initial treatment, nutritional status, psychosocial factors, environmental influences, and drug adverse effects.

For Aim 2, the variables include demographic data, clinical and health history, exposure details, index-TB case data, genetic factors, immunization records, healthcare accessibility, nutritional status, environmental factors, symptom and health monitoring, laboratory results, social and behavioural aspects, contact tracing data, geographic information, and follow-up records.

## Data sources

Datasets for selected TB cohorts will be identified and obtained from the responsible study principal investigators and will comprise digitalized datasets from research studies that recruited index TB cases and household contacts, with follow-up visits at baseline (Month 0), Month 2, and Month 6, and that stored data in electronically accessible formats. Data types expected include demographics, clinical signs and symptoms, treatment regimen and adherence, microbiology (smear/culture/GeneXpert), chest X-ray images, drug susceptibility results, laboratory results, and household contact screening outcomes. The datasets will be reviewed, and a data dictionary will be built prior to machine learning and model evaluation. The purpose of building the dictionary is to facilitate data understanding, feature selection, data pre-processing, and model building, with the ultimate goal of ensuring data quality, facilitating effective model building and evaluation, promoting collaboration, and enhancing the reproducibility of research findings.

## TB treatment cohorts

We shall contact Ugandan principal investigators of approved studies that collected data on TB cases (confirmed and unconfirmed) and their response to treatment, including treatment history and the development of resistance to anti-tuberculosis drugs.

TB index and Contact Tracing Studies

We shall reach out to Ugandan principal investigators who collected data on Index TB cases and their households. These contact-tracing studies investigate the risk of contracting TB and developing resistance to it.

We will enrol only datasets from participants who provided broad consent for the use of their data and stored biospecimens for TB research at enrolment.

## Data management plan

We will adhere to the FAIR (Findable, Interoperable, Accessible, Reusable) principles (ref). Data management will include verifying, organizing, protecting, maintaining, and processing scientific data to ensure it remains accessible, reliable, and high-quality. We will align data management and sharing with FAIR guidelines, leveraging existing data and containers to transform them into machine-readable FAIR Digital Objects (FDOs). All data will be in a digital, machine-understandable format, complete with descriptive metadata, and accessible via a globally unique, persistent, and resolvable identifier. This strategy will improve data integration and visualization across multiple sources, enable granular access controls, and enable decentralized, machine-assisted analysis. We will work with data collectors, managers, analysts, medical professionals, and radiologists to develop ontological models and promote open science practices for existing data. Furthermore, we will generate machine-readable metadata to enhance the findability, accessibility, and reusability of multimodal datasets. These initiatives will support the entire data lifecycle—from data handling and analysis to ML model development—through a cohesive, synergistic approach.

## Data models

We will develop predictive models using three complementary methods: (1) solely clinical tabular data, (2) only chest X-ray imaging data, and (3) a multimodal approach that integrates both modalities. Our aim is to systematically compare unimodal and multimodal strategies to determine the best model configuration that offers improved diagnostic accuracy, robustness, and clinical usefulness. For models based on tabular data, we will thoroughly evaluate various ML algorithms, including Support Vector Machines (SVMs), Random Forests, AdaBoost, XGBoost, Gradient Boosted Trees, and LightGBM. Tree ensemble methods enhance predictive accuracy and robustness by combining multiple decision trees. Boosting algorithms such as AdaBoost, XGBoost, and LightGBM work by sequentially combining weak learners, focusing on misclassified instances at each iteration. Conversely, Random Forests reduce variance and overfitting by aggregating independently trained trees.

For chest X-ray image analysis, we will explore state-of-the-art convolutional neural networks (CNNs) and transformer-based architectures. CNNs excel at extracting hierarchical, spatially coherent features from images, which are then passed to fully connected layers for classification. Transformer-based models complement CNNs by capturing long-range dependencies and contextual information within the images. To mitigate overfitting in image-based models, we will employ comprehensive data augmentation strategies, including rotations, flips, scaling, and intensity variations, to artificially expand the training dataset and enhance model robustness.

## Data Annotation (Labelbox)

The data annotation and quality assurance process begin with uploading raw chest X-ray images to a centralized annotation platform. To ensure diagnostic accuracy and reduce personal bias, each image is assigned to at least two independent annotators who systematically record classification labels, evaluate overall image quality, and create detailed segmentation masks. After completing these initial tasks, the workflow proceeds to a thorough verification phase where outputs from both annotators are compared to identify discrepancies. A series of quality checkpoints act as primary filters to uphold high data standards. During classification and quality checks, the system verifies that both annotations agree and that the image meets the necessary clinical quality standards; failure in either step results in the immediate rejection of the classification label or the entire image. At the same time, segmentation masks are quantitatively evaluated by calculating the Intersection over Union (IoU) between masks created by different annotators. Only masks with an IoU above 0.5 are accepted and combined, while those below are discarded to ensure the spatial grounding used for model training is highly accurate. This structured process produces a “golden” dataset of verified classification labels and segmentation masks, serving as the essential foundation for the model’s spatial and semantic metrics mechanisms. See the process flow in **Figure 1**.

**Figure 1:**
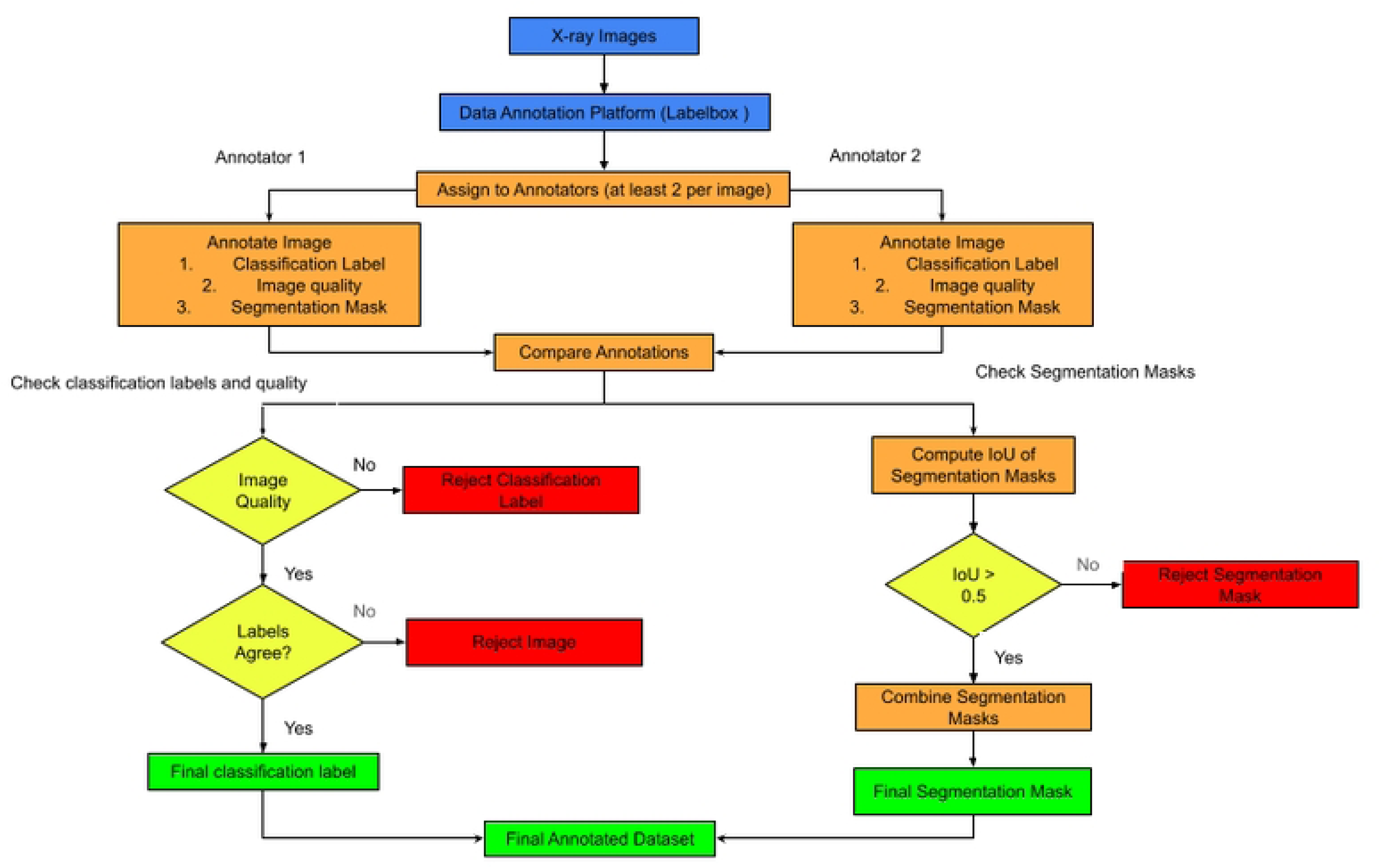
Process flow of Annotating X-ray images using Label box.

## Data Upload Platform (React, Cornerstone)

A platform was developed to enable radiographers to upload digitized X-ray images and assign quality labels. React was used to build the platform’s frontend, while Cornerstone displayed the DICOM images. The system supports two separate data entry workflows based on the user’s professional role. Radiographers ingest medical images, with options for individual file uploads and high-volume batch processing. Once uploaded, images automatically pass through a specialized de-identification pipeline that uses a PHI (Protected Health Information) detection model to identify sensitive data. This data is then masked with black pixels to maintain privacy before the de-identified image is stored. On the other hand, Clinical Officers submit structured data in CSV or Excel formats. After submission, a validation script checks the data’s integrity and format. Successful validation results in data storage and prompts a System Administrator to review the records. If the data fails validation, the upload is rejected, and the officer is immediately notified to correct the issues. Both methods ensure that all stored data is standardized, secure, and ready for clinical use, **see Figure 2**

**Figure 2:**
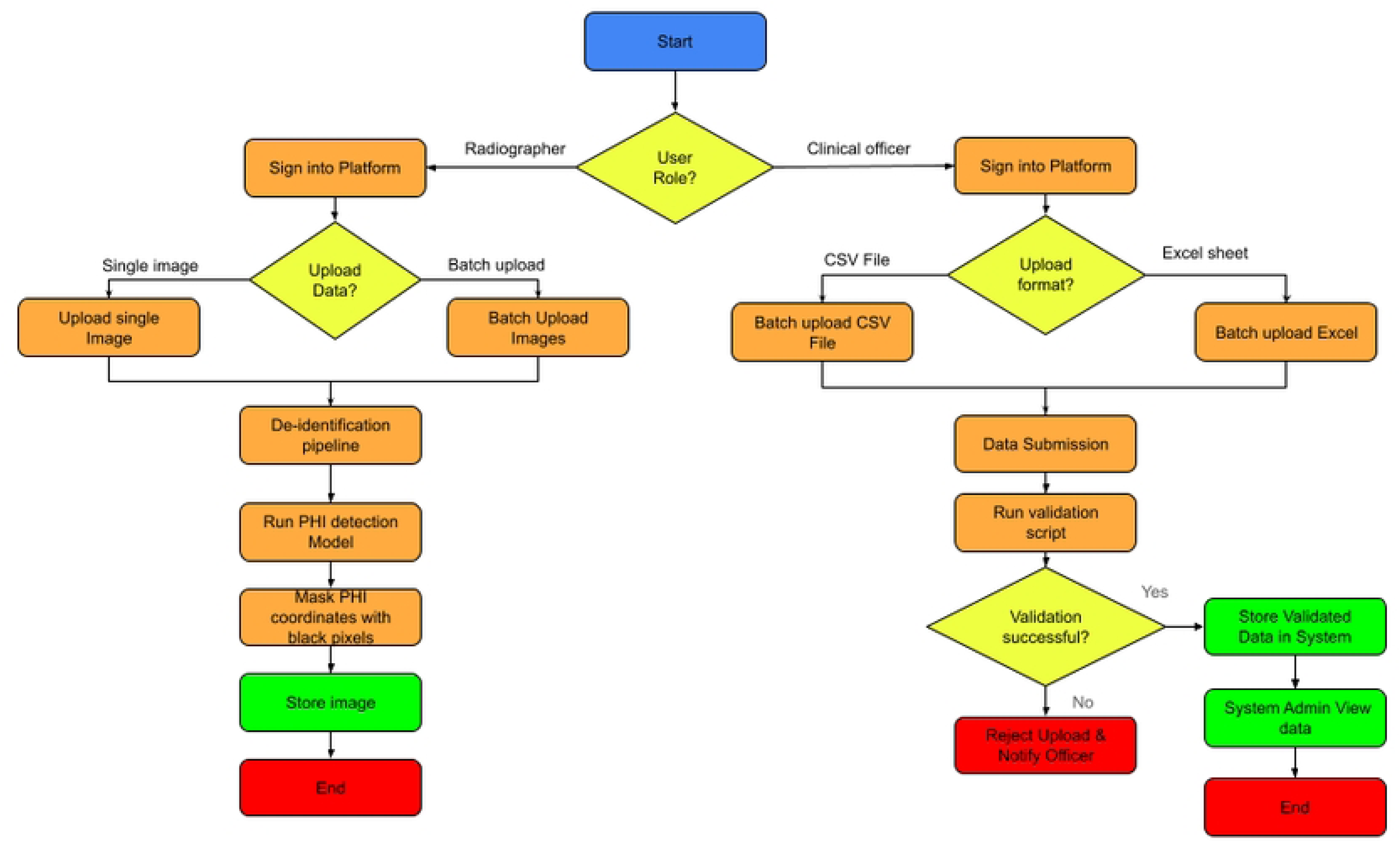
Process flow of data Uploads using React, Cornerstone platform.

## Data Analysis (Pandas, Matplotlib, Seaborn)

The Exploratory Data Analysis (EDA) workflow follows a structured sequence of data-handling and analytical phases to transform raw data into visual insights. The process begins with Authenticated Data Access, ensuring that all data retrieval is performed securely within the system’s permission framework. After successful authentication, the data is ingested and structured using the Pandas library, enabling efficient data reading and manipulation. Once the dataset is prepared, it undergoes rigorous statistical analysis using the Scikit-Learn toolkit to identify underlying patterns, correlations, and anomalies. The final stage of the pipeline involves generating Plot Graphs, which provide a visual representation of the statistical findings to support clinical or technical decision-making, **see Figure 3**.

**Figure 3:**
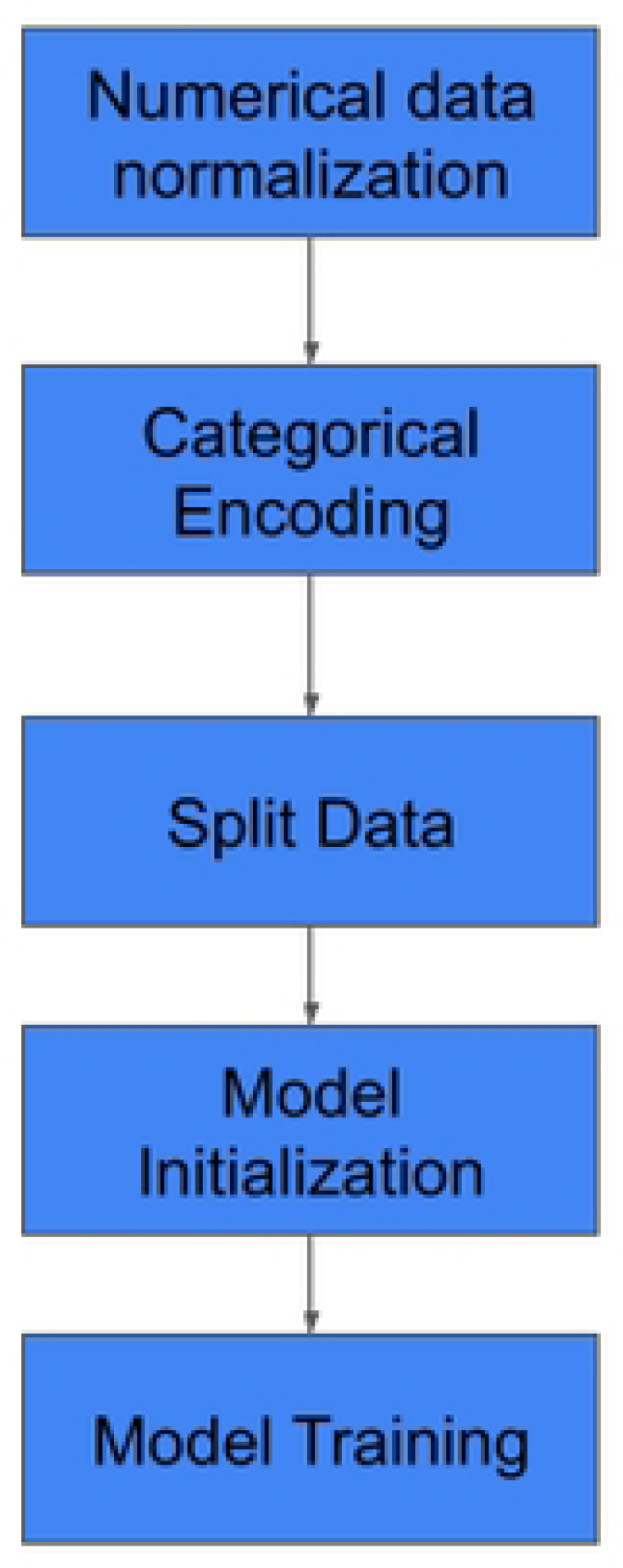
Data Analysis workflow using (Pandas, Matplotlib, Seaborn)

## Model development

The model training process follows a structured sequence that moves from raw data preparation to predictive algorithm deployment, supported by a specialized software ecosystem. The initial phase emphasizes data pre-processing, where raw tabular data is loaded and transformed into a format suitable for ML. This includes numerical data normalization—standardizing values by subtracting the mean and dividing by the standard deviation—and categorical data one-hot encoding, which converts non-numeric variables into numerical vectors. Once the data is refined, the workflow advances to the model training stage using the Scikit-learn library. The processed dataset is first split into a training set (80%) and a testing set (20%) to ensure thorough evaluation. Various ML models, such as the Random Forest Model, are then initialized and trained on the training data to develop predictive models for drug resistance.

To guarantee high performance and reproducibility, the entire training lifecycle is monitored and managed through integrated tools. Weights & Biases is employed for experiment monitoring, offering real-time tracking of model performance and metrics during drug resistance prediction tasks, **see figure 4**.

**Figure 4:**
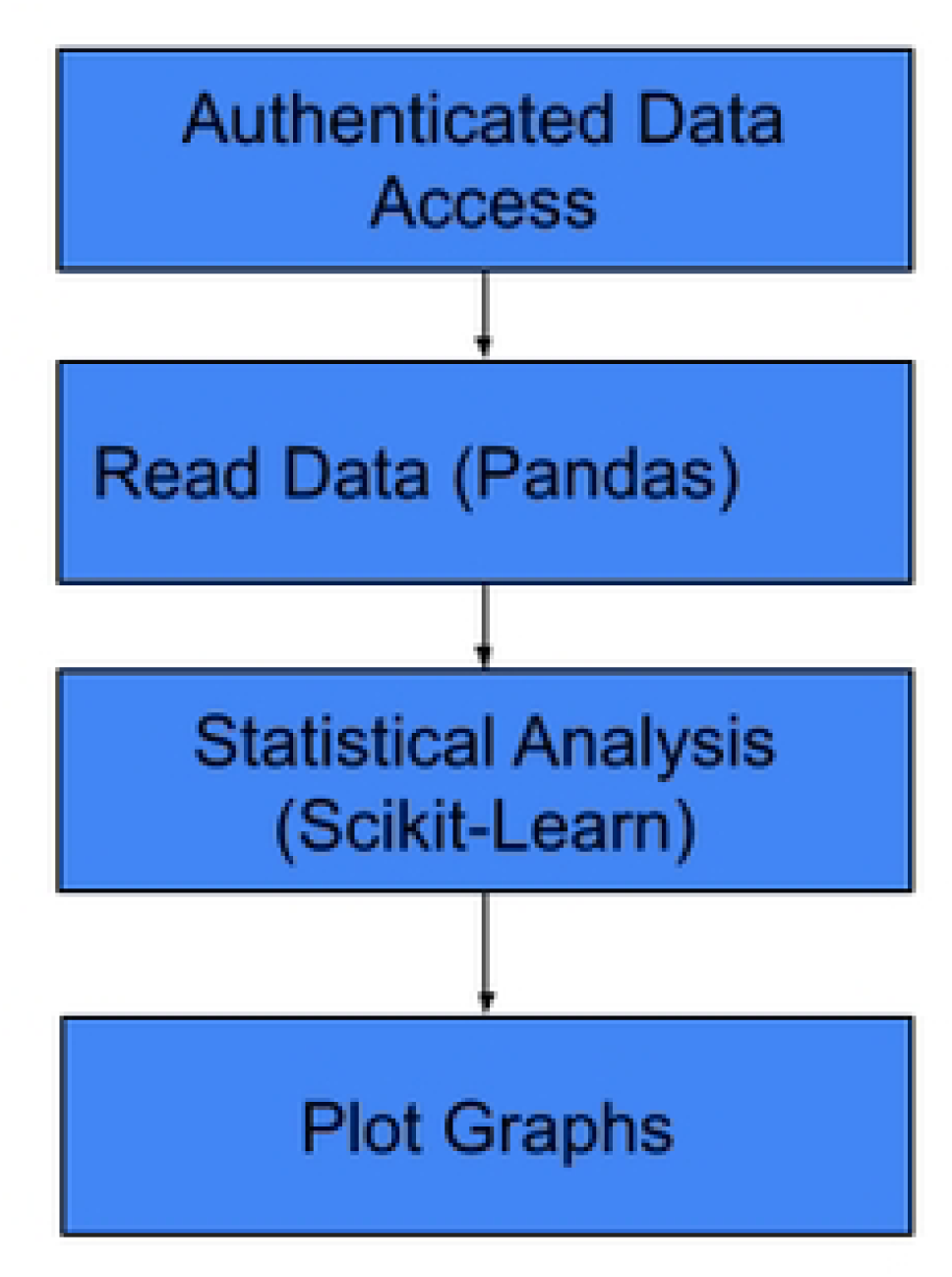
Steps in model development

## Version Control (Git & GitHub)

The software development and codebase are maintained under strict version control using Git, with GitHub serving as the remote repository for collaborative management.

## Ethical approval

The study will be conducted in compliance with the laws and regulatory requirements of Uganda. Local ethical approval was obtained from the School of Biomedical Sciences Research Ethics Committee (SBSREC) and the Uganda National Council for Science and Technology (UNCST) before the study commenced; see reference SBS-2023-495.

## Status and timeline

Identification of research studies with the desired datasets is ongoing. Once a study dataset meets our criteria, we shall obtain administrative approval to use the dataset. This process is ongoing and is expected to be completed in December 2026. Results from the acquired data are expected in March 2027. Throughout this process, the study team will only access de-identified data.

## Discussion

We aim to address the high TB/HIV complexities in high-HIV-prevalence LICs using data-driven approaches to understand the variability of TB phenotypes in Kampala. The study will focus on developing robust, user-friendly data pipelines. The team plans to create AI models with machine learning capabilities to improve TB diagnosis, treatment, and control. The main outcomes include an AI model to predict patients’ responses to TB treatment and another to identify those at risk of drug-resistant TB or likely to develop active disease. Additionally, the project aims to expand these models to forecast disease occurrence, progression, and treatment response for various infectious diseases, illustrating how data-driven tools can support clinical decisions and public health efforts.

## Limitations

We anticipate facing difficulty in obtaining curated data that is AI/ML-readable. However, without addressing these, the use of data science approaches in TB research will remain suboptimal.

## Data Availability

No datasets were generated or analysed during the current study

## Acknowledgements

The authors would like to acknowledge different project Principal Investigators, that is, Drs. Moses Joloba, Willy Ssengooba, and Irene Andia-Biraro, who agreed to share their project datasets. The authors thank Ms. Juliet Kamanyi and Ms. Harriet Nakayiza for the Administrative support.

## Author’s contributions

DPK and EN made substantial contributions to the conception and design of the work; EN and DM wrote the first draft of the manuscript. EN, MT, DPK, AK, RK, FNK, RN, JNN, SN contributed to the final draft. DPK, JNN, EN, and MT reviewed the final draft of the manuscript.

